# SARS-CoV-2 Antibody responses do not predict COVID-19 disease severity

**DOI:** 10.1101/2020.05.15.20103580

**Authors:** William S. Phipps, Jeffrey A. SoRelle, Quan-Zhen Li, Lenin Mahimainathan, Ellen Araj, John Markantonis, Chantale Lacelle, Jyoti Balani, Hiren Parikh, E. Blair Solow, David R. Karp, Ravi Sarode, Alagarraju Muthukumar

## Abstract

**Background:** Initial reports indicate adequate performance of some serological-based SARS-CoV-2 assays. However, additional studies are required to facilitate interpretation of results, including how antibody levels impact immunity and disease course.

**Methods:** In this study, a total of 968 subjects were tested for IgG antibodies reactive to SARS-CoV-2. We confirmed analytic specificity using 656 plasma samples from healthy donors, 49 sera from patients with rheumatic disease, and 90 specimens from individuals positive for PCR-based respiratory viral panel. One-hundred seventy-three cases of confirmed or suspected SARS-CoV-2 were tested for IgG. A subgroup of 37 SARS-CoV-2 PCR-positive cases was tested for nucleocapsid-specific IgM antibody using an in-house developed microarray method. Antibody levels were compared between disease severity groups.

**Results:** All specificity specimens were negative for SARS-CoV-2 IgG antibodies (0/656, 0%). Cross reactivity was not detected in specimens with antinuclear antibodies and rheumatoid factor, or cases with previous diagnosis of viral infection including human coronavirus. Positive agreement of IgG with PCR was 83% of samples confirmed to be more than 14 days from symptom onset, with less than 100% sensitivity attributable to a case with severe immunosuppression. Virus-specific IgM was positive in a higher proportion of cases less than 3 days from symptom onset. No association was observed between mild and severe disease course with respect to IgG and IgM levels.

**Conclusions:** The studied SARS-CoV-2 IgG assay had 100% specificity and no adverse cross-reactivity. Index values of IgG and IgM antibodies did not predict disease severity in our patient population.

## INTRODUCTION

As the COVID-19 global pandemic (1) continues, a major priority is the application of serological testing to determine the scale and rate of exposures. The COVID-19 pathogen,SARS-CoV-2 (2), is an enveloped, positive-sense, single-stranded RNA Betacoronavirus with a ∼30 kilobase genome. The molecular detection of SARS-CoV-2 is based on targeting the viral genome (e.g., Orf1a/b, E, S, N genes) by polymerase chain reaction (PCR) (3–7), and is currently the gold standard to diagnose acute infection (3). Cellular and humoral immunity resolve the infection, which can be detected by the formation of antibodies specific for the virus.

Various serological assays have recently acquired FDA’s emergency use authority (EUA) for SARS-CoV-2 antibody testing in COVID-19 patients, but the interpretation of antibody data and their clinical significance remains challenging. Understanding the time course of antibody response and potential reasons for apparent failure of seroconversion are essential. Further, before assessing whether specific antibodies ameliorate SARS-CoV-2 infection or prevent reinfection, confidence in the analytical specificity of the test is required. Antibody assays are frequently susceptible to non-specific reactivity leading to false positives. This can have dramatic effects when the incidence of exposure is low. Thus, a high positive predictive value gained from minimal cross reactivity towards other pathogen or autoimmune-associated antibodies is critical.

Long, et al. have described a variable antiviral IgM and IgG immune response to SARS-CoV-2 infection in a Chinese population (8), in which seroconversion in a group of 285 patients from 3 hospitals, showed IgG positivity for all cases beyond 17–19 days. Bryan, et al. demonstrated timing of seroconversion for an Idaho population (9). Additional studies are lacking for the U.S. population. The goals of this study were to ascertain key performance metrics of analytical specificity and cross reactivity for a SARS-CoV-2 IgG serological assay, perform a detailed cross-sectional and serial assessment of IgG and IgM antibody responses in suspected COVID-19 patients, and determine their relation to disease severity.

## MATERIALS AND METHODS

### Patient samples

This study was approved by the UT Southwestern Institutional Review Board.A total of 968 individuals (996 total specimens) were included in this study, including 656 healthy controls, 29 patients with systemic lupus erythematosus, 20 with rheumatoid arthritis, 90 with previous positive respiratory viral PCR panel, and 173 confirmed or suspected cases of COVID-19 (**Fig. 1**).

**Figure 1.**
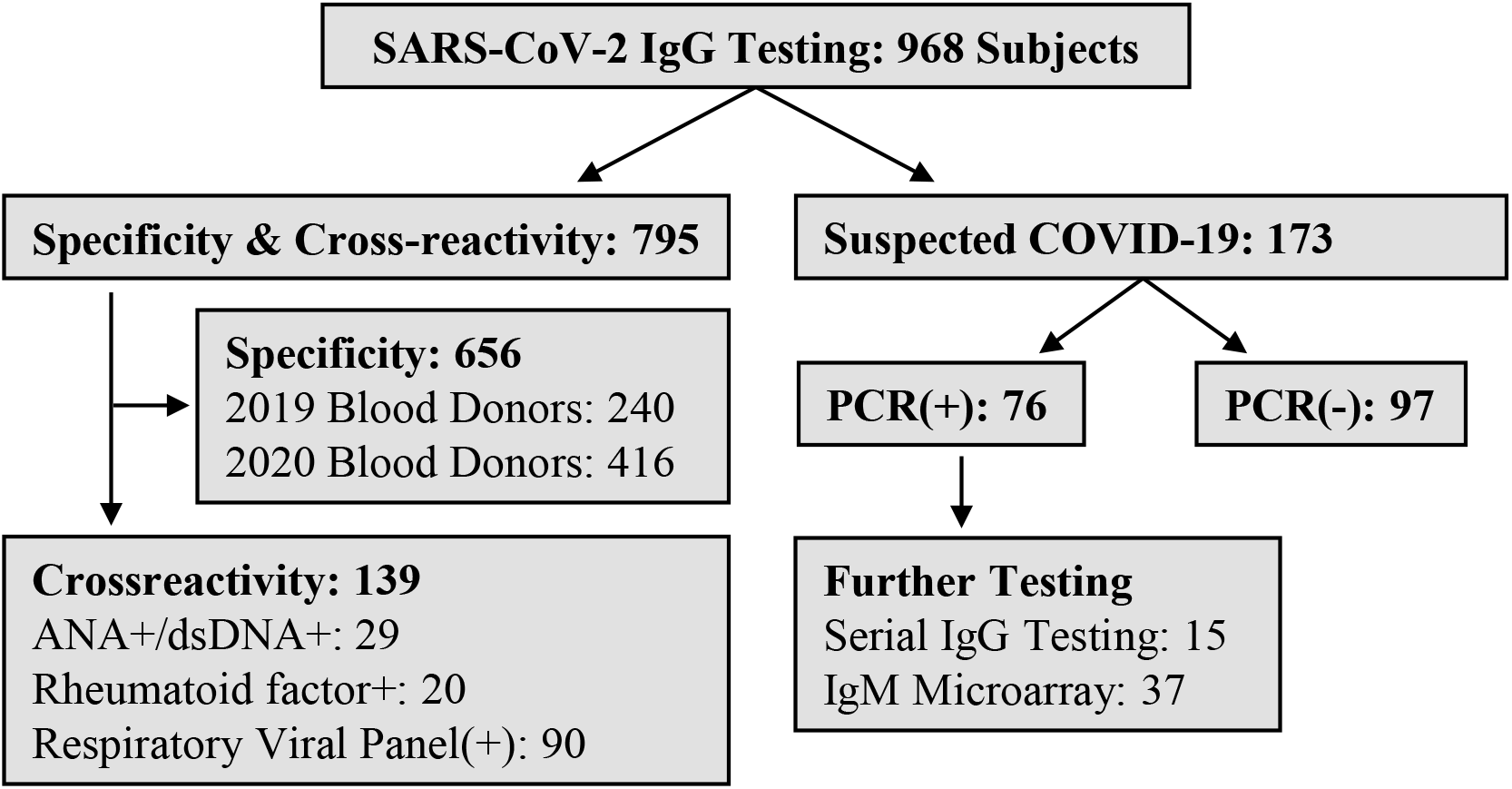
Study Cases. Nine-hundred and sixty eight (968) unique individuals provided samples for SARS-CoV-2 IgG testing, including 15 with serial samples available. IgM testing (not shown) was performed on a group of 37 specimens (17 IgG positive, 20 IgG negative).

### SARS-CoV-2 IgG Testing

SARS-CoV-2 IgG (Abbott 06R86) testing was performed on the Abbott ARCHITECT i2000SR in accordance with manufacturer’s specifications. The test is a chemiluminescent microparticle immunoassay (CMIA) for qualitative detection of IgG antibodies against SARS-CoV-2 nucleocapsid protein (NCP) in human serum and plasma.Strength of response in relative light units (RLU) reflects quantity of IgG present, and is compared to a calibrator to determine the calculated Index (Specimen/Calibrator, S/C) for a sample (with positive at 1.4 or greater).

### SARS-CoV-2 IgM testing

IgM antibody reactivity against SARS-CoV-2 NCP was measured using a laboratory developed protein microarray as described previously (10). Briefly, NCP expressed in baculovirus insect cells (Sino Biological) and in *E. Coli* (Creative Diagnostics) along with control proteins (human IgM and anti-human IgM) were printed onto nitrocellulose membrane coated slides (Grace Bio) in sextuple using a Nanoplotter NP2.1 microarray Inkjet printer (Gesim, Germany). Patient serum samples were diluted 1:100 and incubated with the antigens on the array and the IgM antibody specificities detected with cy5-conjugated anti-human IgM (1:1000, Jackson ImmunoResearch). The array was scanned using Genepix 4400A scanner (Molecular Device) at wavelength 635 nm. The resulting images were analyzed using Genepix Pro 7.0 software (Molecular Devices). The median of the signal intensity for each spot was calculated and the local background around the spot subtracted, and data obtained from sextuple spots were averaged. The background subtracted signal intensity was normalized to the average intensity of the total human IgM (internal positive control) to generate normalized signal intensity (NSI). Samples with NSI ≥25 were considered positive for IgM. The NSI of NCP IgM was used to generate heat maps using Cluster and Treeview software (http://bonsai.hgc.jp/~mdehoon/software/cluster/index.html).

### Analytical specificity

Specificity was evaluated using 240 banked plasma samples collected prior to the COVID-19 pandemic (blood donors September through November 2019), and an additional 416 healthy donors without recent illness collected from March to April, 2020.

### Cross-reactivity studies

Cross reactivity specimens were collected by cross referencing banked serum in the HLA lab (January 1, 2015- September 30, 2019) with patients who had previously tested positive for cytomegalovirus (CMV IgG), influenza A/B, RSV, or an endemic Coronavirus (NL63, 229E, OC43 or HKU1) by viral molecular tests. As the patients may have been immunosuppressed, we included only those specimens having normal or high levels of total IgG (measured alongside SARS-CoV-2) with no infusion of intravenous immunoglobulin in the preceding 3 months. Interfering substance specimens came from a collection of residual serum from a study of systemic lupus erythematosus patients that were positive for ANA and other autoantibodies (n= 29 collected 2004–2007). Patient samples strongly positive for rheumatoid factor (n= 20 collected 2011–2014) were also evaluated.

### Agreement with PCR-based testing

Agreement wih PCR-based molecular testing was determined using 173 plasma samples collected (147 lithium heparin, 13 EDTA, 12 sodium citrate, and 1 sodium fluoride anticoagulants) from suspected COVID-19 cases with prior or same-day PCR-based nasopharyngeal swab testing on the m2000 Abbott RealTi*m*e SARS Cov-2 assay or the Abbott ID NOW^™^ COVID-19 assay. Patient charts were reviewed to determine time between symptom onset (fever, respiratory symptoms, or gastrointestinal complaints) and severity of condition (whether or not intensive care was required). A subgroup of 37 PCR-positive cases (17 IgG positive, 20 IgG negative) were additionally evaluated for SARS-CoV-2 IgM.

### Serial Patient Monitoring

For 15 PCR-positive cases, two to six serial measurements were performed using available residual plasma samples. IgG levels and seroconversion based on calculated Index (S/C) were tracked over time.

### Statistics

The calculated Index (Specimen/Calibrator, S/C, IgG level) was provided by the instrument. When multiple values of IgG S/C were compared, a mean and standard deviation were calculated. Student’s t-test was used to compare two groups of non-parametrically distributed data and p-value <0.05 was considered significant.

## RESULTS

### Analytical Specificity

The SARS-CoV-2 IgG assay was calibrated followed by an imprecision study performed over a period of 5 consecutive days and was found to be acceptable. Analytical specificity of the assay was evaluated with samples from healthy blood donors and none of these samples (0/656) were positive for virus-specific IgG (**Table 1**) and the mean index value was 0.04, well below the cut-off of 1.4 for a positive index value.

**Table 1.** SARS-CoV-2 IgG Results in Healthy Donors and Cases of Previous Respiratory Viral Infection.

| Sample Type | IgG, AU/mL |
| --- | --- |
| **Blood Donors** |  |
| Mean±SD | 0.039±0.065 |
| Positive of n tests (%) | 0/656 (0%) |
| **CMV IgG+** |  |
| Mean±SD | 0.07±0.067 |
| n of Positive tests (%) | 0/23 (0%) |
| **Flu A+** |  |
| Mean±SD | 0.13±0.19 |
| n of Positive tests (%) | 0/8 (0%) |
| **Flu B+** |  |
| Mean±SD | 0.11±0.17 |
| n of Positive tests (%) | 0/7 (0%) |
| **RSV+** |  |
| Mean±SD | 0.035±0.029 |
| n of Positive tests (%) | 0/6 (0%) |
| **Coronavirus +** |  |
| Mean±SD | 0.050±0.079 |
| n of Positive tests (%) | 0/47 (0%) |

### Cross-reactivity studies

To determine whether antibodies formed in response to viral respiratory infections may cross-react with the SARS-CoV-2 antigen (NCP) on the assay’s paramagnetic beads, we included samples of patients who had tested positive on a molecular based respiratory viral panel test (**Table 1**). We excluded any patients treated with intravenous immunoglobulin in the last 3 months. As some patients were post-transplant and on immunosuppression medications, we quantitated total IgG and excluded any samples with hypogammaglobulinemia (low IgG) to reduce false negative results. We tested 23 CMV IgG positive samples and none were COVID-19 IgG positive (0/23, 0%). No cases associated with prior Flu A+ (n=8), Flu B+ (n=7), RSV+ (n=6), or all 4 types of human coronavirus (n=47) demonstrated cross-reactivity (0/90, 0%).

Likewise, we tested 29 samples from lupus patients that were positive for multiple autoantibodies (100% ANA, 62% anti-dsDNA, 75% anti-U1RNP, 55% anti-Sm, 34% anti-Ro52, and 24% anti-La) and an additional 20 samples from rheumatoid arthritis patients positive for rheumatoid factor (85% were also anti-CCP positive). None of these sera with clinically significant levels of autoantibodies produced a positive antiviral IgG test result (0/49, 0%) (**Table 2**). Highest mean of S/C ratio observed was 0.05 for human coronaviruses and 0.08 for rheumatic diseases.

**Table 2.** Autoantibody Interference.

| Autoantibody | IgG Positivity Rate (%) |
| --- | --- |
| **Systemic lupus erythematosus (29 Patients)<sup>a</sup>** |  |
| ANA | 0/29 (0%) |
| Anti-DNA | 0/17 (0%) |
| Anti-U1RNP | 0/21 (0%) |
| Anti-Sm | 0/15 (0%) |
| Anti-Ro52 | 0/10 (0%) |
| Anti-La | 0/7 (0%) |
| **Rheumatoid arthritis (20 Patients)<sup>b</sup>** |  |
| Rheumatoid factor (RF) | 0/20 (0%) |
| Anti-CCP | 0/17 (0%) |

<sup>a</sup> Cases (29 total) were positive for 1 or more of the autoantibodies listed.
<sup>b</sup> All cases were RF positive, with 17/20 (85%) also anti-CCP positive.

### Cross-Sectional data for SARS-CoV-2 IgG and IgM

Of 173 suspected COVID-19 cases, 76 were confirmed positive by PCR methods. Overall, 29 of 76 (38%) tested positive for SARS-CoV-2 IgG. The time course of symptom onset revealed increasing IgG positivity rates (**Table 3**) from <3 days (1/15, 7%), to 3–7 days (8/27, 30%), 5–15 (5/15, 33%), and being the highest after 14 days (5/6, 83%). IgG positivity was high (10/13, 77%) for patients with indeterminate time from symptom onset. IgM testing (**Fig. 2**) performed on 37 PCR positive specimens showed positivity in 9 of 17 (53%) IgG positive cases and, interestingly, in 7 of 20 (35%) IgG negative samples. IgM positivity occurred at larger proportion for <3 days (3/6, 50%) compared to IgG, but at similar rates overall at days 3–7 (4/11, 36%), days 8–13 (4/11, 36%), and after 2 weeks(4/5, 80%). IgM positivity was low (1/4, 25%) for patients with indeterminate time from symptom onset.

**Table 3.** SARS-CoV-2 IgG Positive Agreement by Days Post-Symptom Onset.

| Time from symptom onset * | IgG Positivity Rate (%) | IgM Positivity Rate (%) |
| --- | --- | --- |
| <3 days | 1/15 (7%) | 3/6 (50%) |
| 3-7 days | 8/27 (30%) | 4/11 (36%) |
| 8-13 days | 5/15 (33%) | 4/11(36%) |
| >14 days | 5/6 (83%) | 4/5 (80%) |
| Indeterminate | 10/13 (77%) | 1/4 (25%) |
| **Total** | **29/76 (38%)** | **16/37 (43%)** |

\* For RT-PCR confirmed SARS-CoV-2 cases

**Figure 2.**
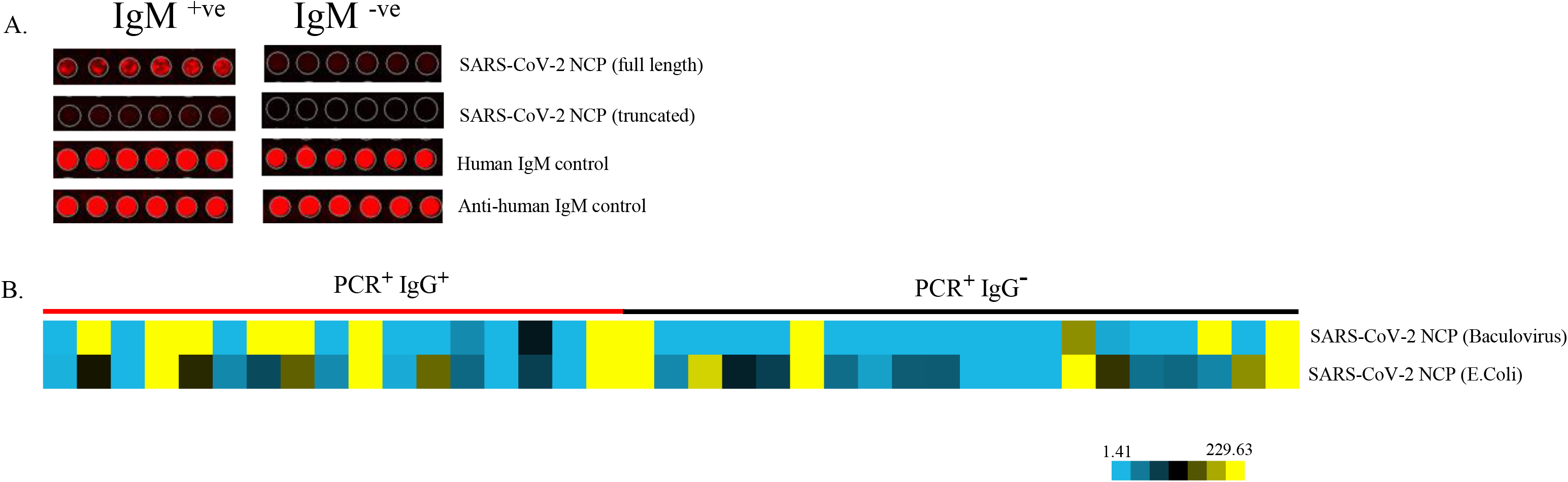
IgM Microarray Analysis. Array images of IgM positive and negative samples are shown (A) as well as heatmap of IgM anti-SARS-CoV-2 NCP are shown for IgG positive and negative cases of confirmed COVID-19.

SARS-CoV-2 IgG antibody results agreed with the PCR negative samples for 96 of 97 (99%) of cases, including 55 instances of patients with new or acute-on-chronic symptoms suspicious for COVID-19 and with known time of onset.

### Disease severity and IgG and IgM value

We hypothesized that a more severe disease course was related to an increased immune response, which may result in a higher level of SARS-CoV-2 IgG antibody reactivity. Cytokine storm has been implicated as a potential life-threatening event in SARS-CoV-2 infection, and this would activate many aspects of the immune system including the humoral antibody response. We compared IgG levels from all SARS-CoV-2 PCR-positive patients who had a mild/ moderate disease course to those who had severe disease (admitted to the ICU), and there was no difference in IgG antibody levels between the two groups (**Fig 3A**).

**Figure 3.**
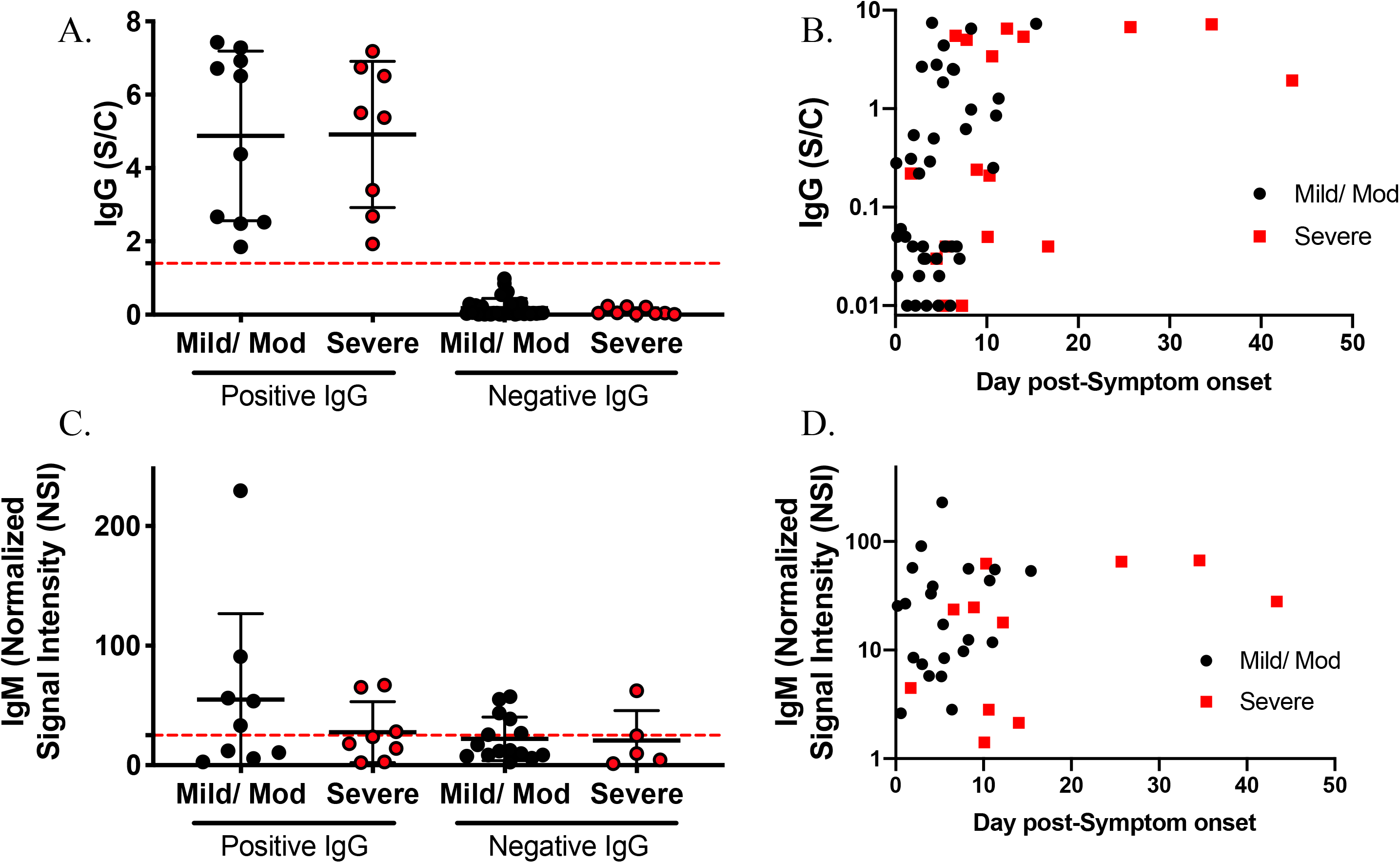
Antibody levels by disease severity for PCR+ subjects. (**A**) SARS-CoV-2 specific IgG antibody results which were positive or negative were divided by disease severity and (**B**) plotted against number of days from symptom onset. (**C**) SARS-CoV-2 nucleocapsid specific IgM antibody results were divided by IgG positivity to demonstrate when a sample was IgM+, but IgG−. (**D**) IgM antibody levels were plotted against number of days from symptom onset. Middle line is the mean bars represent standard deviation. Black dots are mild/ moderate cases, while red dots represent severe cases. The red dash line in (B) represents the negative cut off level.

Next, we assessed the impact of timing of collection on the antibody response by comparing the number of days since symptom onset between mild/ moderate and severe disease status (**Fig. 3B**). Severely affected patients had higher IgG antibody levels measured at a later time compared to mild cases (p<0.05). Similarly, higher IgM levels were observed in severely affected patients (**Fig. 3C, D**). It is possible that the course of IgG levels was qualitatively different in severe patients, so data from serially collected IgG samples was plotted against day of symptom onset(**Fig. 4**). Severely affected patients were tracked longer, because they were hospitalized longer, but a similar early increase in antibody titers was observed in mild/moderately affected patients when compared to severely affected patients. Interestingly, one patient was seronegative even on day 28, but this was attributed to immunosuppression to prevent cardiac transplant rejection.

**Figure 4.**
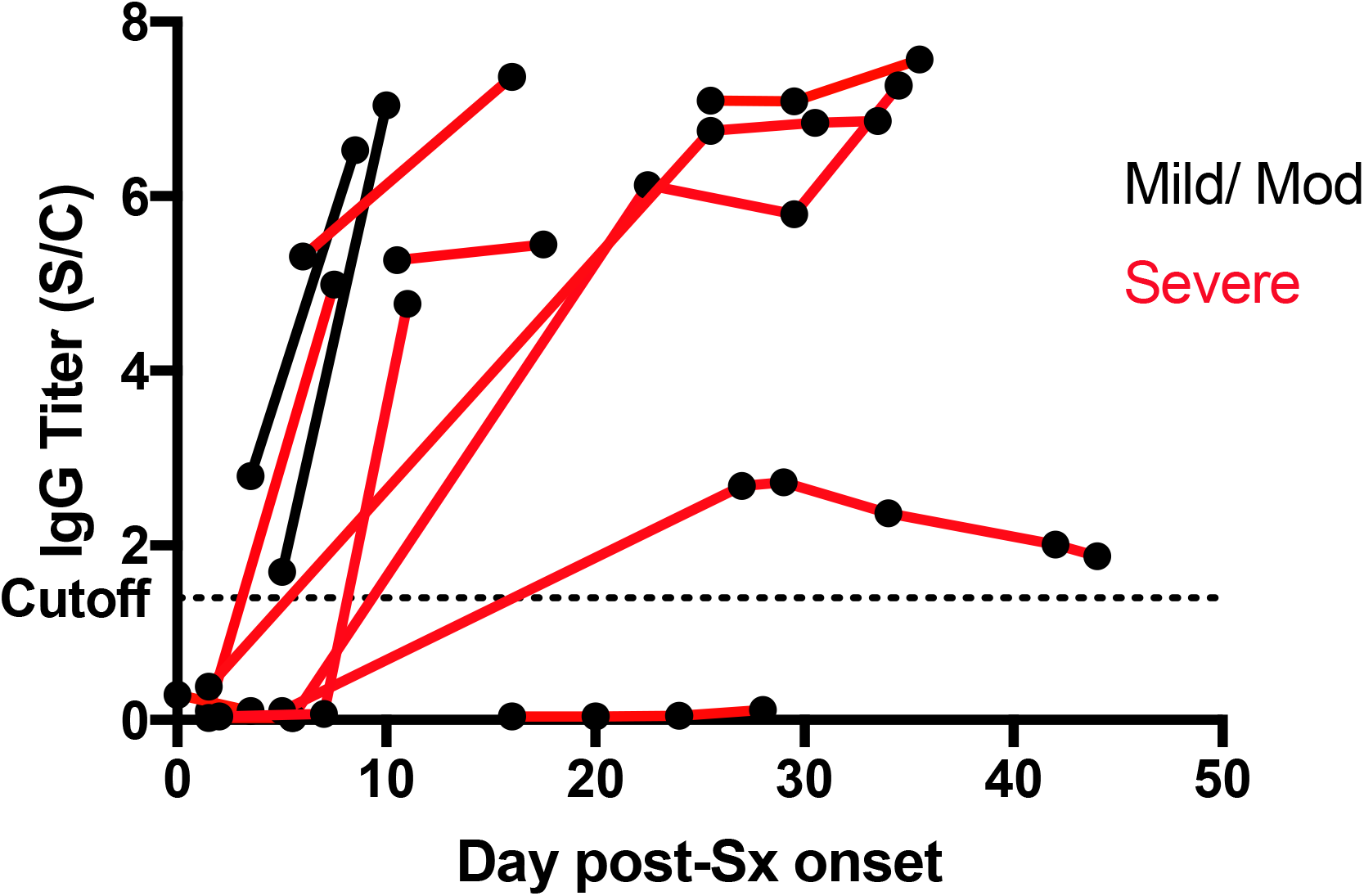
Serial IgG measurements. For patients with multiple samples taken, the IgG level was plotted against time from symptom onset. Black dots represent the IgG level at a specific time. Samples from the same patient are connected by either red (severe cases) or black lines

### Serial Patient Monitoring and Seroconversion

Thirty-eight samples were available from 13 patients with known date of symptom onset (Fig. 3C) and 4 samples from 2 patients with indeterminate date of symptom onset. Within this group, 77% (10/13) became IgG positive, including specifically 0% (0/8) for less than 3 days post symptom onset, 33% (3/9) at 3–7 days post-symptom onset, 86% (6/7) at 8–13 days post-symptom onset, and 91% (10/11) at more than 14 days (**Figure 4**). For those where seroconversion was not observed, samples were only available for less than 7 days from symptom onset for 2 cases or patient was subject to significant immunosuppression. For the two cases with indeterminate date of symptom onset, one demonstrated seroconversion between samples 11 days apart. The second case did not demonstrate seroconversion over 9 days.

## DISCUSSION

Here we confirmed the high specificity reported by the manufacturer for a SARS-CoV-2 IgG serological assay, using comparatively larger groups for certain rheumatological conditions and infections. Notably, CMV IgG did not cause assay interference despite potential false positivity reported by the manufacturer. Rheumatoid factor is an anti-human antibody (IgM or IgG) that, if complexed with other human immunoglobulins, could falsely increase positivity of an assay. However, we observed no interference by rheumatoid factor in 20 samples. Testing 47 samples with prior endemic coronavirus infection yielded no false positives. Negative agreement between IgG and PCR indicated only one case testing IgG positive despite negative PCR testing. This initial PCR result was later determined to be a false negative based on evaluation using an alternative molecular platform. Positive agreement with PCR was lower in the early stages of infection, increasing with time from symptom onset, yet not as quickly compared to the manufacturer’s report.

Overall, our results largely corroborate and add to the findings by Bryan, et al. (9) who evaluating the same platform. The study showed high specificity in testing 1020 specimens submitted for HSV Western blot serology from before the COVID-19 pandemic. As such, specificity and cross reactivity were not specifically addressed in the setting of underlying rheumatologic disease or previous endemic coronavirus. A possible difference between our findings was sensitivity after 14 days of symptoms. In our study, a single negative case attributed to a patient with marked immunosuppression resulted in reduced sensitivity beyond 14 days. An unknown factor in similar studies are the number of cases included with severe underlying immunosuppression. For instance, a recent publication by Long, et al. also indicated 100% IgG positivity at 17–19 days. This latter study utilized a different assay and focused on a population in China, and was thus not as comparable. However, the same question persists regarding the makeup of comorbidities in the test population and highlights that discrepancies may arise in antibody response when comparing serology in unequal groups. Nonetheless, within our serial testing group, given the higher number of cases beyond 14 days, we did encounter sensitivity of 91% (10/11), which was closer to the findings of Bryan and Long. As with Bryan, et al., we have noticed alternative cutoff values for IgG level could be utilized with potentially beneficial diagnostic effects. As an example, lowering the cutoff by half (to 0.7) would capture an additional four cases with mid-range days from symptom onset (5 to 11) without any loss in specificity based the PCR result.

Long, et al. had reported a counter-intuitive peak of IgM positivity (20–22 days) later than for IgG positivity (17–19 days). IgM responses usually peak within the first week after infection and before IgG class switching. When early in infection, IgG may not yet be positive. When we tested samples for IgM reactivity, seven IgG negative cases were positive for IgM. These samples were positive for IgM earlier than IgG, with onset ranging from 0 to 11 days from symptom onset. This increased the sensitivity by 35% within the IgG negative samples tested (7/20) and improved diagnostic utility by 9% overall (7/76).

As described, we segregated our IgG and IgM results based on severity (ICU care versus no ICU care). Long, et al. indicated that a severe disease course resulted in a high IgG level during the second week of disease that becomes indistinguishable from milder cases after 14 days (9). We did not observe such a difference using a different CMIA method. This could be due to fewer patient samples, but the significance of their finding was very strong, which indicates it should have replicated were it a real phenomenon. IgM levels in our study showed no significant difference when analyzed by disease severity. Thus, antibody levels themselves do not appear to reflect disease severity.

A major hurdle to validation was access to patients after a sufficient time period of infection because most patients presented before 14 days from symptom onset. Limited resources and self-quarantine measures have impaired repeated testing for serial testing at a later date.Consequently, less data on mild and moderate patients existed compared to patients admitted to the ICU. We do however have the advantage of reviewing medical charts to find examples of false negative by PCR and false negative by serology. These examples indicate that molecular and serologic testing have complementary roles in tracking exposure to SARS-CoV-2. Our data does not provide information on how long IgG stays positive in the long term or whether it specifically confers immunity.

## CONCLUSIONS

As communities continue to grapple with the COVID-19 pandemic, reliable measures of previous exposure and immunity are essential. Several platforms are now coming into broader clinical use, providing a window into the SARS-CoV-2 antibody response. Widespread efforts to track SARS-CoV-2 patients for antibody development will clarify expectations for when testing should return positive, situations in which seroconversion may fail, and what the antibody response can tell us in patients with active infections.

## Data Availability

The authors confirm that the data supporting the findings of this study are available within the article [and/or] its supplementary material.

## ACKNOWLEDGMENTS

We would like to thank Clements University Hospital Core lab staff, especially Brittany Diaz,Sylynn Garza and Charles Alexis for serology testing and archiving of all the samples related to this study. We also acknowledge Carter Blood Care (Bedford, TX) for their archived patient samples for specificity studies and the Human Leukocyte Antigen lab at UT Southwestern for their help in retrieving banked serum samples for cross-reactivity studies, as well as Dr. Ashley Young for aid in chart review.

## Commercial Interest Disclosure

The following authors have no interests to disclose: WP, JS, QL, LM, EA, JM, CL, JB, HP, EBS,DK, RS, AM

The following authors have these interests to disclose: None

